# Five years follow-up of patients with non-melanoma skin cancer treated with HeberFERON

**DOI:** 10.1101/2022.02.07.22270604

**Authors:** Y Duncan-Roberts, Y Garcia-Vega, S Collazo-Caballero, M Rodriguez-Garcia, M Zalazar-Sedano, JL Rodríguez-Rojas, A Tuero-Iglesias, C Valenzuela-Silva, I Raices-Cruz, N Castro-Basart, E Garcia-Iglesias, L Pereda-Lamela, R Hernández-Rodríguez, E Arteaga-Hernandez, V Muzio-Gonzalez, I Bello-Rivero

**Affiliations:** Department of Clinical Investigations, Center for Genetic Engineering and Biotechnology, Ave 31 e/ 158 and 190. Cubanacan, Playa, 10600, Havana, Cuba

**Keywords:** Basal cell carcinoma, squamous cell carcinoma, interferons, HeberFERON

## Abstract

**Introduction:** Non-melanoma skin cancer is the most common tumor. The combination of IFN-alpha 2b and IFN-gamma has been used as a new therapeutic opportunity to treat basal cell carcinomas and cutaneous squamous cell carcinomas. The aim of this report is to record prospectively the recurrence and new lesions rates in patients participating in phase II clinical trials.

**Methods:** Phase II clinical trials (double-blind randomized one center study, InCarbacel-III, in patients with basal cell carcinoma; and open, non-randomized multicenter study, CECIN, in patients with cutaneous squamous cell carcinomas), with the use of the combination of IFN-alpha 2b and IFN-gamma were conducted to evaluate the efficacy, safety and the 5-year duration of clinical responses. Both studies were approved by institutional ethic committees and all the patients given their written informed consent. The investigational treatment was administered, peri- or intralesionally, three times per week, during 3 weeks. Clinical (RECIST 1.0) responses were evaluated three months after the end of treatment.

**Results:** The combination of IFNs in InCarbacel-III study showed the best clinical response (complete response of 64.3%, overall response of 85.7%) with the highest doses (10.5 MIU); without patient’s recurrence at 5 years follow-up (3.5 MUI and 10.5 MUI groups). The frequency of new lesions decreased in the treated patients 8 times. In the CECIN study 14 patients achieved complete response and 4 partial responses (overall response rate 67%). Up to the 5-year follow-up none of the patients with complete response had recurrence or new lesion. In both studies the cosmetic results were excellent and the reported adverse events were mostly of mild intensity.

**Conclusions:** The use of the combination of IFN-alpha 2b and IFN-gamma showed efficacy in basal cell carcinoma and cutaneous squamous cell carcinoma promoting a long term response for at least 5 years and decreasing the rate of new lesions, safely and with excellent cosmetic effects.

## Introduction

Non-melanoma skin cancers (NMSCs) are the most common forms of human neoplasia. The annual incidence of NMSCs is estimated to be higher than 5.4 million cases in USA^1^, most of which are unreported. Basal cell carcinomas (BCC) and cutaneous squamous cell carcinomas (CSCC), referred to as keratinocyte carcinoma (KC), represent approximately between 80% and 20 % of all NMSC cases, respectively^2,3^. However, likely, the ageing of population is influencing in an increase of CSCC cases, comparable with BCC^1^. BCC and CSCC account for approximately more than 6000 case in male population, with more than 5000 females cases annually, in Cuba^4^. BCC and CSCC have a favorable prognosis with surgical resection, when detected in the early stages and their metastatic potential is low.

BCC is a slow growing tumor with invasion and destruction of adjacent tissues. Five-year recurrence rate in BCC is variable, depending of the treatment used, 3% [electrodesiccation and curettage (ED&C)]^5^, and 15% to 39% (cryosurgery)^6,7^ Five-year recurrence rates for radiation therapy of BCC are between 4% and 16%^8^. A 10-year recurrence rate for primary BCC was 12.2% using standard surgical excision and 4.4% for Mohs micrographic surgery (MMS). The 10-year recurrence rate for recurrent facial BCC was 13.5% for standard surgical excision and 3.9% for MMS^9,10^. Metastasis is very rare in BCC (0.0028% to 0.55%).^11^ More than 90% of CSCCs are cured by initial treatment. Recurrence of CSCC has been found to 0.8% and 1.7% for cryosurgery and ED&C, respectively, in small, low-risk tumors^12^. Data from studies yielded a 26.4% odds of recurrence following PDT^13^. After surgical excision or MMS, a 5.4% and 3% recurrence rate has been observed, respectivelly^12^. The 5-year recurrence rate is 8% and the 5-year rate of metastasis is 5%^14^.

A definitive management strategy for the treatment of advanced NMSC has not been established. Over the past decades several therapies, including Hedgehog inhibitors for BCC, monoclonal antibodies targeting epidermal growth factor receptor (EGFR), cetuximab and panitumumab; and immune check-point, pembrolizumab for CSCC, has been approved or are in study^13,15^. Resistance to these inhibitors has been identified^16,17,18,19^. Then, new treatment options are needed to obtain specific advantages over current approaches that can offer therapeutic opportunities for non-responders and relapses.

Recently a new formulation of IFNs that combine IFN-alpha 2b and IFN-gamma (HeberPAG/HeberFERON) is being showing encouraging therapeutic and cosmetic effect in NMSC^20,21,22,23, 24, 25^

The aim of this report is the evaluation of the 5-year recurrence and new lesions rates in patients with BCC or CSCC treated with HeberFERON (IFN-alpha 2b and IFN-gamma) during phase II clinical trials.

## Methods

### Study design and patients

The trials were designed jointly by the principal investigator and the clinical trial monitors of the Center for Genetic Engineering and Biotechnology (CIGB). Data were collected by the institution investigators participating in the studies, under a confidentiality agreement and were retained and analyzed by the sponsor.

Patients, both genders, older than 18 years, who gave their written informed consent to participate, were enrolled in the trials. The studies excluded pregnant or nursing women, patients with known hypersensitivity to IFN or with history of autoimmune diseases. Clinical (RECIST 1.1)^26^ responses were evaluated.

InCarbacel III study (code: IG/ IAIIGI/NB/0601; registroclinico.sld.cu/en/trials/RPCEC00000066) was a double blind, randomized, controlled, one center phase II trial, of dose evaluation (group A: 0.75×10^6^ IU IFN-alpha 2b and 0.125×10^6^ IU IFN-gamma; group B: 1.5×10^6^ IU IFN-alpha 2b and 0.25×10^6^ IU IFN-gamma; group C: 3.0×10^6^ IU IFN-alpha 2b and 0.5×10^6^ IU IFN-gamma; group D: 6.0×10^6^ IU IFN-alpha 2b and 1.0×10^6^ IU IFN-gamma; and group E: 9.0×10^6^ IU IFN-alpha 2b and 1.5×10^6^ IU IFN-gamma), conducted in histological confirmed BCC patients, of any subtype, or localization, size 1.0-5.0 cm.

CECIN study (code: IG/AGI/NE/0901, registroclinico.sld.cu/en/trials/RPCEC00000144) was an open, non-randomized multicenter study in patients with CSCC (stage I and II), between 1.5 and 5.0 cm of infiltrative clinical subtype treated with 10×10^6^ IU IFN-alpha 2b and 1.5×10^6^ IU IFN-gamma.

The trial protocols were approved by the Ethics Committee and the Scientific Review Board of participating institutions (Hermanos Ameijeiras, Havana and Amalia Simoni, Camaguey, Hospitals in Cuba) in accordance with the ethical principles stated in the Declaration of Helsinki.

### Procedures

All the patients were treated in the medical oncological/dermatological certified cabinets in participating health centers. Before treatment, each patient had a medical history, a physical and skin examination for disease assessment, documentation of concurrent medications, a punch biopsy of not more than 25% of the total lesion size.

A single lesion per patients was treated with HeberFERON. In the case of patients with more than one lesion, the tumor to be treated was selected as per size or the lesion localized nearest to the others ones. HeberFERON was administered by perilesional/intratumoral routes, 3 times per week during 3 consecutive weeks, on an outpatient basis in certified hospital cabinets. Only the treated lesions were evaluated for their characteristic and clinical response to the treatment. Lesion diameter (d) measurements were done using a Folding Magnifier, rulers or calipers, systematically during the treatment time and until week final evaluation and photographed for documentation.

Clinical response was categorized as complete response (CR)^26^ defined as no residual BCC or CSCC; partial response (PR) as ≥30% reduction in tumor size and stabile diseases (SD) as < 30% reduction in the tumor size; and progression (P) by increase in the lesion size.

Safety was monitored by adverse events control and their frequency calculated. Additionally, blood samples were taken for routine hematological and biochemical determinations.

Subjects were examined as outpatients. Patients’ follow-up examinations were done at weeks 1, 2, 3 and 12-14 after treatment onset, with final evaluation (clinical, laboratory tests) were done at 12-14 weeks. Patients with CR or PR were fallowed during five years for tumor recurrences and/or appearance of new lesions of the same type of tumor. The case report forms (CRF) were filled by principal or responsible investigators or designed specialists. The data were monitored and then imported by double entry in electronic base data and validated and processed on Microsoft Access and then imported to SPSS version 13.0 for analysis by the independent statistic group at department of clinical investigations of the sponsor (CIGB).

The frequency distributions for qualitative variables and central tendency and dispersion were estimated: mean, standard deviation, median, interquartile range (OR), maximum and

For each type of adverse event, were estimated frequency distributions (IC95%) with classical statistic. The laboratory data were analyzed as a paired (initial-final result) result using paired T student and Wilkinson tests, depending from Shapiro-Wilk test results.

### Outcomes

The primary endpoint was the percentage of CR after one cycle of administration (3 weeks of treatment) and a follow for final response evaluation of 12-14 weeks from the end of treatment. The secondary endpoints were the demographic characteristics of patients and the characteristics of BCC or CSCC lesions, safety and recurrence and new lesions rates.

## Results

During the InCarbacel-III study were enrolled 75 patients with BCC, average 61.5 years–old with predominance of males (54.1%), where 89.2% were white. Skin phototype II and III were predominant, 65.3% presented one lesion, the median size of them was 2.4 cm and disease stage I was the most frequent. The lesions were more common localized in the trunk and the superficial clinical subtype was the most represented (Table 1).

**Table 1.**
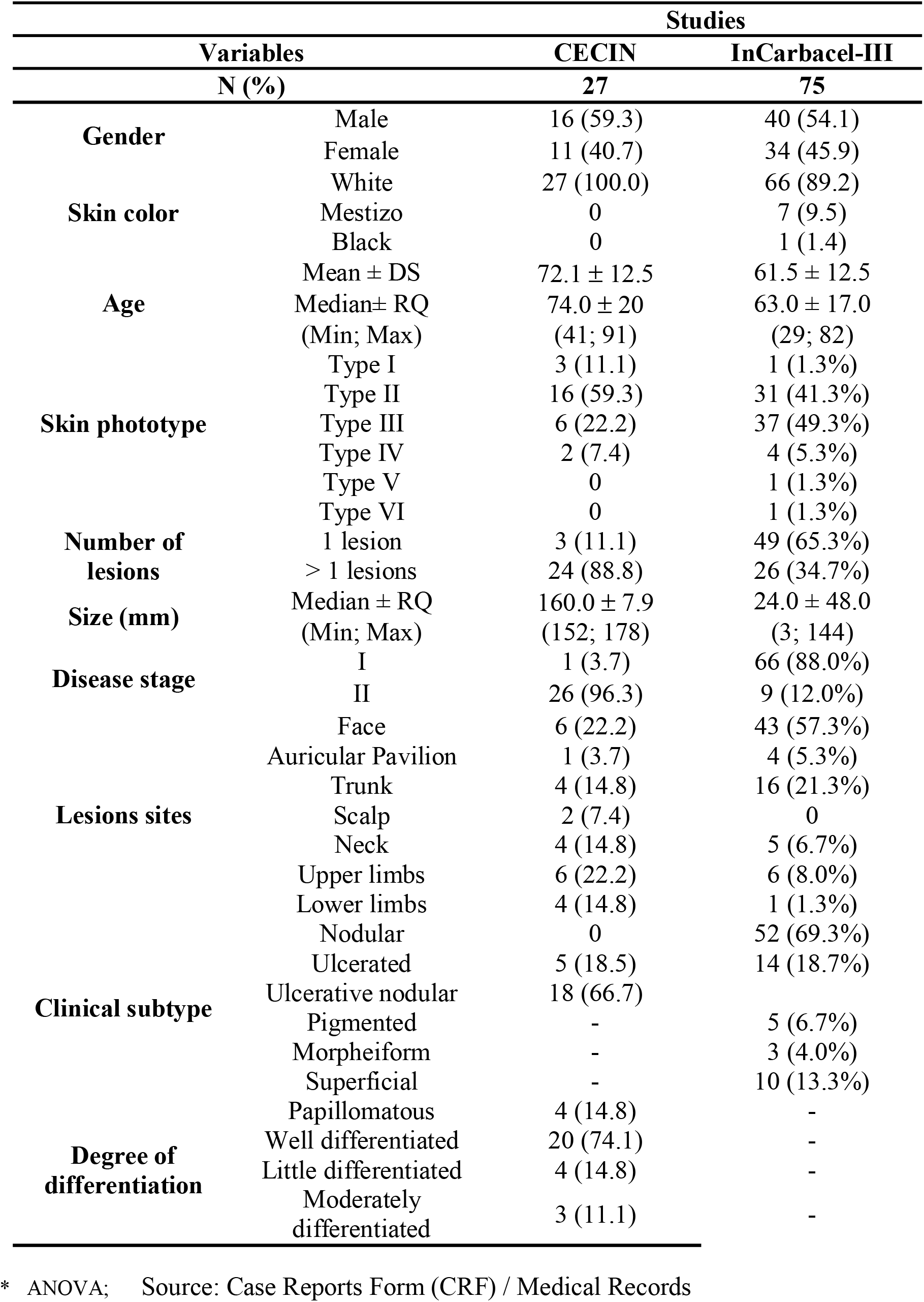
Baseline characteristics and demographics of patients from InCarbacel-III and CECIN studies.

Table 2a shows the efficacy of the treatments in the study InCarbacel-III. Clinically, by intention to treat (ITT) analysis, the groups with higher HeberFERON doses (D and E), had the highest CRs, 60% and 64%, respectively. The aesthetic outcome was evaluated in patients with CR (figure 1). Overall, the quality of healing was good; behaving similarly in all groups, being 100% good in groups C and E.

**Table 2a.** Clinical response of patients with BCC treated with HeberFERON. InCarbacel-III Study.

| Clinical Response | Group A | Group B | Group C | Group D | Group E | Total | FB (P(H<sub>0</sub>)) |
| --- | --- | --- | --- | --- | --- | --- | --- |
| N (%) | 14 | 15 | 14 | 15 | 14 | 72 |  |
| CR | 6 (43) | 8 (53.3) | 8 (57.1) | 9 (60.0) | 9 (64.3) | 40 (56.0) | 0.048(0.95) |
| PR | 4 (26) | 5 (33) | 3 (24) | 5 (33) | 3 (24) | 20 (28) |  |
| SD | 4 (26) | 2 (13.3) | 3 (21.4) | 1 (6.7) | 2 (14.3) | 12 (16.7) |  |
| ORR | 10 (71) | 13 (87) | 11 (79) | 14 (93) | 12 (86) | 60 (83) | 0.106(0.90) |
| $P[Resp > 0.75/$ | 0.221 | 0.705 | 0.421 | 0.962 | 0.820 | 0.221 | |

**Figure 1.**
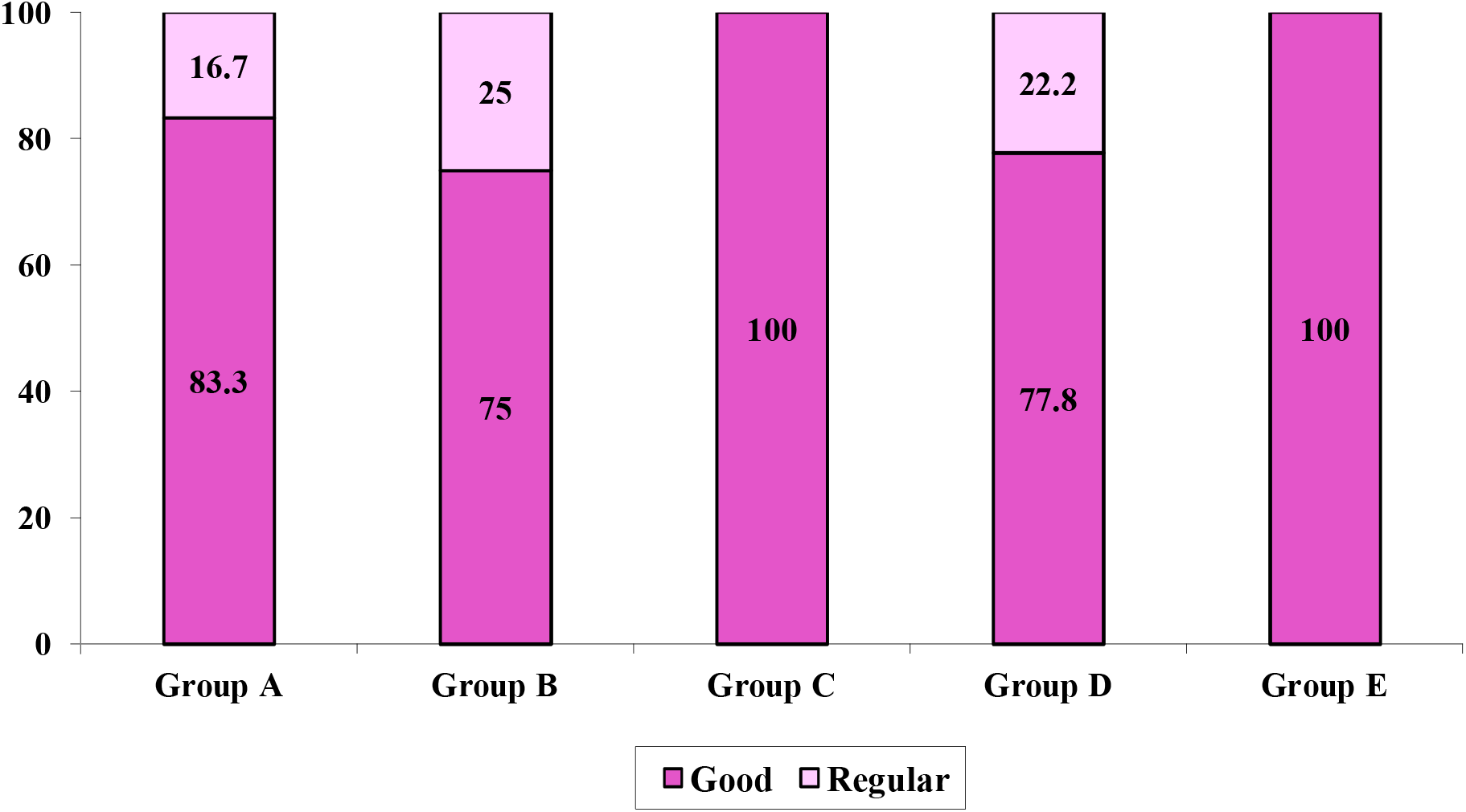
Cosmetic results of patients with CR in the InCarbacel-III study. The groups C and D showed the best cosmetic results.

Adverse events (AEs) occurred in 96% of the patients with BCC, with fever, chills, headache, arthralgia, malaise, asthenia, perilesional edema and erythema, anorexia, nausea, decreased white blood cells, diarrhea, dry mouth and drowsiness, being reported more frequently (> 10%). Of the total of 1072 AEs presented in the patients under study, it was observed that 93.0% were of mild intensity, 6.5% moderate and only 0.5 % serious. A causal relationship was established as highly probable, for 97.1% of events.

The analysis of risk vs benefit was done considering as benefic the number of CR (40 out of 75 patients) and as risk, the occurrence of moderate or severe AEs (75 events out of 1072). The result of the analysis produced a Bayes factor (FB) >1, indicting a benefit over risk in all the groups of treatment.

Twenty-seven patients with CSCC, average 72.1 years–old, predominantly male (59.3%) initiated the treatment in the CECIN study, with predominance of skin phototype II with 89% bearing more than one skin lesion. The average size of lesions was 16 cm, and 96.3% were in stage II. The lesions were localized mainly in the face and upper limbs (22.2% of patients, respectively). Ulcerative nodular clinical subtype was commonest and 74% were well differentiated (Table 1).

Fourteen patients (51.9%) had CR, 4 (14.8%) PR, 8 (29.6%) had stable disease (EE) and 1 patient (3.7 %) was not evaluable (NE). The ORR was of 66.7%. The aesthetic evaluation in patients with CR or PR found 77.8% of good quality of healing (see table 2b).

**Table 2b.** Clinical response of patients with CSCC treated with HeberFERON. CECIN study.

| Clinical Response | Total | FB (P(H<sub>0</sub>)) |
| --- | --- | --- |
| N (%) | 27 |  |
| CR | 14 (51.9) | (31.2; 72.6) |
| PR | 4 (14.8) | (4.2; 33.7) |
| SD | 8(29.6) | (10.6; 48.7) |
| NE | 1 (3.7) | (0.1; 19.0) |
| ORR | 18 (66.7) | (47.0; 86.3) |
| NR | 9 (33.3) | (13.7; 53.0) |

ITT: Intention to treat; CR: Complete response; PR: Partial response; SD: Stable disease; ORR (CR+PR): Overall response rate.

The AEs with a higher frequency (≥10%) were fever (55%), general malaise (40%), chills (33%), headache (18%), myalgia (18%), anorexia (14%), asthenia (11%) and arthralgia (11%). Overall there were 100 AEs, 82.0% of mild intensity, 17.0% moderate and 1.0% of severe intensity. 62.0% of the events were classified as possible and 16.0% as probable.

### Five year follow-up

Forty-six patients with BCC (61.3%) distributed in the five treatment groups were evaluated in the 5 years follow-up, as shown in Table 3, of them the 36 with CR and 10 patients with PR. Of the 36 patients evaluated during these 5 years with CR, 3 (8.3%) had reappearance of the treated lesion: Group A – 2/3 (66.6%), Group B – 1/7 (14.3%), and Group D – 1/5 (20%). None of patients from groups C and E had recurrences. The followed patients from groups A, C and E did not develops new BCC lesions (Table 4). Overall, the rate of new lesions of BCC in patients treated with HeberFERON was 8.0%.

**Table 3.** Recurrences rate in non-melanoma skin cancer patients treated with HeberFERON. Five years follow-up.

|  | InCarbacel-III study |  |  |  |  | Total |
| --- | --- | --- | --- | --- | --- | --- |
|  | A | B | C | D | E |  |
| Enrolled patients | 15 | 16 | 15 | 15 | 14 | 75 |
| CR per group | 6 | 8 | 6 | 8 | 8 | 36 |
| Year of follow-up | Followed/# recurrences |  |  |  |  |  |
| 1 | 6/0 | 8/0 | 6/0 | 8/0 | 8/0 | 36/0 |
| 2 | 6/1 | 8/0 | 5/0 | 7/0 | 7/0 | 33/1 |
| 3 | 4/0 | 8/0 | 4/0 | 7/1 | 7/0 | 30/1 |
| 4 | 1/0 | 8/1 | 3/0 | 5/0 | 5/0 | 22/1 |
| 5 | 3/2 | 7/1 | 6/0 | 5/1 | 7/0 | 28/4 |
| Recurrence rate at 5-years | 66.6% | 14.3% | 0% | 20% | 0% | 14.3% |
| CECIN study |  |  |  |  |  |  |
| Patients with CR | 14 |  |  |  |  |  |
| Year of follow-up | Followed/# recurrences |  |  |  |  |  |
| 1 | 14/0 |  |  |  |  |  |
| 2 | 14/0 |  |  |  |  |  |
| 3 | 11/0 |  |  |  |  |  |
| 4 | 8/0 |  |  |  |  |  |
| 5 | 8/0 |  |  |  |  |  |
| Recurrence rate at 5-years | 0% |  |  |  |  |  |

**Table 4.**
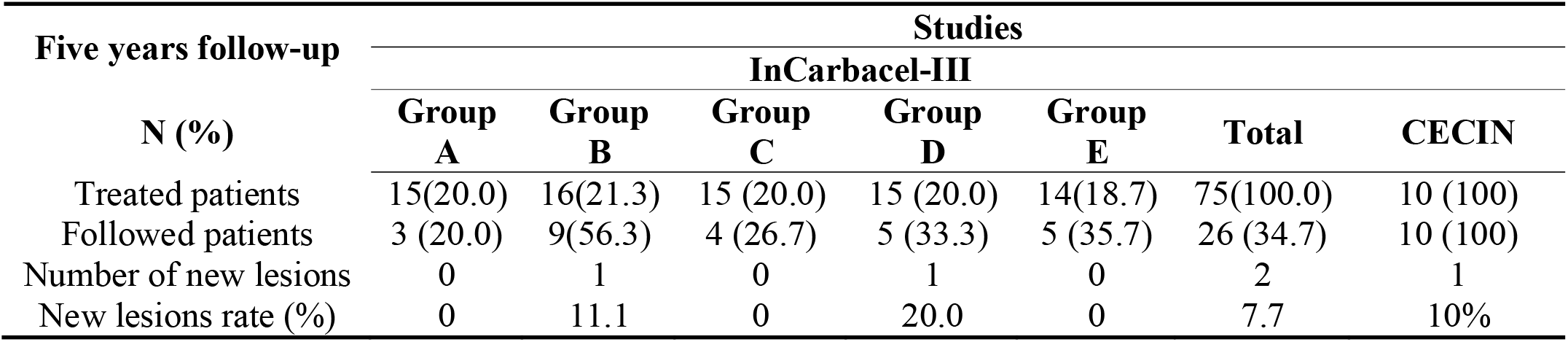
The rate of new lesion in non-melanoma skin cancer patients treated with HeberFERON. Five years follow-up.

Eight patients (80%) with CSCC were also followed for 5 years to see the occurrence of recurrence or appearance of new lesions (8 with CR, and 2 with PR). None of the evaluated patients had a recurrence and only one had a new lesion.

## Discussion

IFNs are indicated in the treatment of several neoplasms among which are BCC and CSCC^27^. In ours studies the clinical response was good supported by the clinical and cosmetic results. The results obtained indicate that the treatment with the HeberFERON is a highly favorable clinical intervention to avoid the tumor recurrences. In the case of BCC, the patients belonging to groups C (3.5 MIU) and E (10.5 MIU) were those who did not have recurrences of the treated lesions after five years follow-up. In the CSCC patients (treated with 11.5 MIU), no recurrence were observed at all in the followed patients.

This may be due to the fact that after exposure to interferons, there is an increase in the number and activity of cytotoxic cell lymphocytes, which determines an additional increase in immune status^28,29,30,31,32^.

The observed antitumor effect could be related to the increase in apoptosis via Fas-FasL^33^ and with the induction of the expression of the tumor suppressor gene, p53^34^. It could also be a consequence of the antiproliferative and immunomodulatory effects of IFNs ^35,36^.

The reduction in the frequency of the appearance of new lesions in patients with BCC is another element to be highlighted as impact for this treatment. It has been described that approximately 25% of patients with BCC can develop a new lesion^37^. Overall, this treatment reduces this probability approximately 3 times, over the course of 5 years. No new lesions were detected after 5 years follow-up in in some patients. In the case of patients with CSCC none of patient developed a new lesion. In neither of the two entities are there references to the use of a similar product that allows comparisons to be made.

The treatment with HeberFERON was effective, doses dependent and safe in patients with non-melanoma skin cancer. The aesthetic results are very encouraging. No keloids formation was observed. Confirmation of these results in a larger cohort of patients in a phase III clinical trials comparing with recent approved immunotherapies and/or target therapies in patients with BCC and CSCC is recommended.

## Data Availability

Qualified individuals may request access to the de-identified participant data, anonymized clinical study reports, informed consent forms, through submission of a proposal with a defined research question to the corresponding author, Bello-Rivero Iraldo, provided that the necessary data protection and ethical committee approvals are in compliance with the trial. An agreement for transfer of these data will be required.

## Declarations

Study execution conformed to the ethical principles of the Declaration of Helsinki and the International Council for Harmonization of Good Clinical Practice guidelines. The authors were responsible for designing the trial and for collecting and analyzing the data.

## Authors’ contributions

YDR and YGV contributed to the design of the protocol, supervised the clinical trial execution, results interpretation and contribute to the writing of the manuscript. SCC, MRG, MZS, JLRR were the clinical investigators and supervised treatment of patients in the clinical practice, and recorded the information in medical records of the patients included from the clinical site in their provinces. ATI, CVS, IRC, EGI were the statistics of the trial design and data processing. NCB was responsible for laboratory test. LPL and RHR compiled the data and verified the quality of data. EAH conducted the histological evaluation for tumor diagnosis. VMG supervised and assessor the trials. IBR contributed to the design of trials protocols, supervised the trial, interpreted and discussed all the data and results, and contributed to the writing of the manuscript. All authors read and approved the final manuscript. All authors read and approved the final manuscript.

## Acknowledgements

We thank the specialists and technical staff from the Dermatological Services, and Histological Studies from the health institution participating in the studies. We also acknowledge personnel that carried out and were responsible for logistic assurances, randomization of trials investigational drugs, and theirs distribution.

